# Cancer Medicine Prices, Availability, and Affordability in Kisumu County, Western Kenya

**DOI:** 10.64898/2026.05.27.26354206

**Authors:** James Onyuro Oketch, Stephen Asito Amolo, Daniel Ogungu Onguru

## Abstract

**Background:** The rising prices of cancer medicines have intensified concerns about treatment access and health system sustainability particularly in low- and middle-income settings. Facility level evidence on what medicines is actually available, at what prices, and at what cost to patients remains scarce, constraining evidence-based policy reform.

**Methods:** Using adapted WHO/Health action international methodology, we conducted a cross-sectional survey of 52 cancer medicines across five therapeutic classes at five health facilities in Kisumu County, Kenya. Availability was measured as the proportion of facilities stocking each medicine. Affordability was assessed using days’ wages required for the lowest-paid government worker to purchase standard treatment regimens.

**Results:** Overall medicine availability was 48.1%. The breast cancer AC regimen required 19.6 to 47.4 days’ wages per full course; cervical cancer cisplatin, 19.8 to 49.2 days’ wages, colorectal FOLFOX, 80.0 to 303.6 days’ wages, and prostate docetaxel reached 437 days’ wages at the highest-cost facility. The Social Health Authority’s (SHA) KES 550,000 annual ceiling adequately covered cytotoxic regimens for common cancers at competitive prices but was exceeded by 24 to 116% for HER2-positive breast cancer requiring trastuzumab, with further strain for recurrent cervical and metastatic prostate cancers. A subsequent cap to KES 800,000, substantially improves adequacy for standard cytotoxic chemotherapy but does not resolve gaps for HER2-targeted or continuous hormonal therapies.

**Conclusions:** Cancer medicines in Kisumu County are inconsistently available and highly variable in price. We call for urgent retail price markup regulation, expanded pooled procurement, inclusion of priority targeted therapies on the Kenya Essential Medicines List, and SHA benefit packages redesigned around full-course regimen costs.

**HIGHLIGHTS:**

- Only 48% of essential cancer medicines were available in Kisumu.
- Full-course chemotherapy can cost patients up to 437 days’ wages.
- Kenya’s insurance cap inadequately covers patients needing targeted therapies.
- Universal health coverage cannot succeed without medicine price reform.

## 1. BACKGROUND

Cancer is a leading cause of death worldwide, accounting for approximately one in six deaths globally, with incidence projected to exceed 28 million cases annually by 2040 (1). The burden falls disproportionately on low- and middle-income countries (LMICs), where over half of all cancer deaths occur and health systems are least equipped to respond. In sub-Saharan Africa, the cancer burden is rapidly increasing alongside rising demand for effective, safe, and affordable treatment. In Kenya, cancer represents a major public health crisis, with over 44,726 new cases and nearly 30,000 deaths annually (2, 3). Breast, cervical, prostate, oesophageal, and colorectal malignancies are the most common, all requiring systemic pharmacological treatment for meaningful clinical outcomes. Access to essential cancer medicines is therefore a critical determinant of survival, reduction in disease-related illness, and premature death.

Despite their clinical centrality, cancer medicines remain poorly accessible across many LMICs. Approximately 40% of essential cancer medicines listed by the WHO are available to patients only at full out-of-pocket cost (4). Evidence shows that cancer medicine prices continue to increase globally, often outpacing national health budgets and household incomes (5). In sub-Saharan Africa, access is eroded by high medicine prices, fragmented supply chains, and the notable presence of substandard or falsified oncology products infiltrating the market (6–8). A systematic scoping review across the WHO African region identified major subnational evidence gaps in medicine availability data and called for robust facility-level evidence to guide targeted policy interventions within African health systems (9).

Existing studies across the region illustrate the scale of the problem. In Kenya, Rwanda, and Uganda, several WHO-listed essential cancer medicines are largely unavailable or unaffordable in both public and private facilities (8). Treating early-stage breast cancer across Cameroon, Ethiopia, Kenya, and Malawi has been estimated to cost the equivalent of 47–242 days’ minimum wages, and colorectal cancer 233–869 days’ wages (7). These figures point to the near-impossibility of treatment for patients without insurance. In Ghana, a World health organization/health action international-based survey (WHO/HAI) found substantial price variability across facility types, with generic medicines dominating available products and several essential oncology medicines either absent or rarely stocked (10). In Kenya specifically, chemotherapy availability for childhood cancers at major referral hospitals has been reported at only 48.5%, with frequent stock-outs of methotrexate, bleomycin, etoposide, and ifosfamide forcing treatment interruptions (11).

A major barrier to addressing these gaps is the limited transparency in pharmaceutical pricing, facility-level data on medicine prices and availability (12). Without such data, governments cannot conduct effective forecasting, budgetary planning, or price negotiation. Although national monitoring systems exist, comparatively less attention has been given to inter-facility price differences or the real patient-level cost of completing treatment. Consequently, the design and implementation of targeted interventions, including price regulation, procurement reform, and financial protection strategies remain difficult.

In Kenya, the Social Health Authority (SHA) increased the annual oncology benefit package to Kenya Shillings (KES) 550,000 per patient, removing prior limits on chemotherapy cycles within the annual cap (13). Whether this package is adequate for full-course treatment of common cancers remains uncertain, given the cumulative costs of chemotherapy, targeted therapy, diagnostics, and supportive medicines. Evidence from Rwanda demonstrates that insurance improvements alone do not eliminate patient financial barriers when individual medicine costs remain high (14). Redesigning benefit packages requires granular, facility-level cost data. Therefore, this study assessed cancer medicine availability, inter-facility price variability, and regimen-based affordability across public and private health facilities in Kisumu County, Kenya.

## 2. METHODS

### 2.1 Study Design and Setting

This was a facility-based cross-sectional study conducted in October 2025, using a standardized and locally adapted WHO/HAI methodology to assess medicine prices, availability, and affordability of essential cancer medicines in Kisumu County, Kenya. In addition to standard WHO/HAI indicators, the study incorporated regimen-based costing using national treatment protocols and analysis of inter-facility price variation to enhance policy relevance. The study adapted the WHO/HAI framework by replacing MPR analysis with inter-facility range ratios and days’ wages affordability, due to absence of reliable international reference price data for the full medicine list. All medicine prices reflect patient selling prices recorded during this survey period. Medicine prices were recorded in Kenyan Shillings (KES) and converted to US dollars (USD) using the exchange rate prevailing at the time of data collection (KES 129.50 = USD 1.00). The study was conducted across five facilities including one major public referral hospital, one major private hospital, and three private clinics located within a 2 km radius. These facilities constitute the main urban cancer care cluster in Kisumu County and represent the primary dispensing points for oncology medicines, reflecting typical patient care pathways.

### 2.2 Study Population and Sampling

The study population comprised medicine outlets (hospital pharmacies and private clinics) involved in dispensing cancer medicines. A census approach included all five eligible facilities within the study area, consistent with the WHO/HAI minimum recommendation of five or more outlets per sector. Cancer medicines were selected based on the Kenya Essential Medicines List (KEML, 2023) and commonly prescribed non-KEML medicines. Both originator brands and lowest-priced generic (LPG) equivalents were assessed where available. A preliminary spot check on wholesalers and private chemists within Kisumu established that three major wholesalers stocked approximately three to four or no cancer medicines, and were therefore excluded.

### 2.3 Data Collection

Data was collected using a structured questionnaire adapted from the WHO/HAI standard data collection tool. Key methodological modifications included: (i) inclusion of chemotherapy regimen costing based on national treatment protocols (2019), and (ii) collection of inter-facility patient-price variation to inform procurement benchmarking. Availability was determined through physical verification of stock on the day of the facility visit. Prices were recorded as final patient prices excluding additional service charges to ensure comparability (15). Data collection was supervised and reviewed daily by the principal investigator to ensure completeness and accuracy. Double data entry and 20% random verification were applied. Any inconsistencies were resolved through follow-up with the relevant pharmacists.

### 2.4 Key Definitions and Calculations

Availability was defined as the percentage of medicine outlets where a specific medicine was physically present on the day of the survey. Medicine prices were recorded in KES and converted to USD at KES 129.50 per USD to enable international comparison.

Affordability was calculated as the number of days’ wages required for the lowest-paid government worker to purchase a specific medicine dose or regimen cycle, using the Government of Kenya minimum wage of KES 732.44 (USD 5.65) per day.

Overall availability was calculated as the total number of medicines available (stocked) across all facilities and all medicine types divided by the total number of possible availability instances (52 medicines × 5 facilities = 260), expressed as a percentage.

Category-level availability was calculated using the same method but restricted to the medicines within each therapeutic class.

Regimen costs were estimated using BSA = 1.7 m² and Kenya’s 2019 national oncology treatment protocols. Full vials were costed at listed patient prices with no pro-rating for partial vial use. Where the calculated dose exceeded a single vial, the minimum number of whole vials required were costed per regimen. Full-course estimates used the minimum number of cycles for best-case and maximum for worst-case scenarios.

### 2.5 Data Analysis

Data was entered and analyzed using a modified WHO/HAI Medicine Price and Availability Workbook (Microsoft Excel V15.0). Descriptive statistics (means, medians, frequencies, and percentages) were used to summarize availability and price data across sectors. Affordability was expressed in days’ wages. Inter-facility price variability was assessed using price range ratios separately for generic and originator brand products.

### 2.6 Ethical Considerations

This study involved human participants (health facility pharmacists) who provided medicine price and availability data. Ethical approval was obtained from the Jaramogi Oginga Odinga Teaching and Referral Hospital Institutional Scientific and Ethics Review Committee (JOOTRH-ISERC; approval number ISERC/JOOTRH/128/2025). A research license was obtained from the National Commission for Science, Technology and Innovation (NACOSTI; license number NACOSTI/P/25/4176229), with institutional and county-level authorizations from the Jaramogi Oginga Odinga University of Science and Technology Board of Postgraduate Studies (Ref. H162/P/6241/23) and the Kisumu County Government Department of Medical Services, Public Health and Sanitation (Ref. GN 133 VOL.XXI/(72)). Written informed consent was obtained from all participants. Confidentiality was ensured through unique identifiers, password-protected data storage, and aggregate reporting to prevent identification of individual facilities or respondents.

## 3. RESULTS

### 3.1 Overall and Category-Level Availability

A total of 52 cancer medicines across five therapeutic classes were assessed in five health facilities. Overall availability was 48.1% across all surveyed facilities and all medicine types. Mean availability varied substantially across therapeutic classes. Supportive medicines showed the highest availability (65%), followed by cytotoxic medicines (63.1%), hormonal therapies (53%), targeted therapies (20%), and immunosuppressive medicines (10%) (Figure 1).

**Figure 1:**
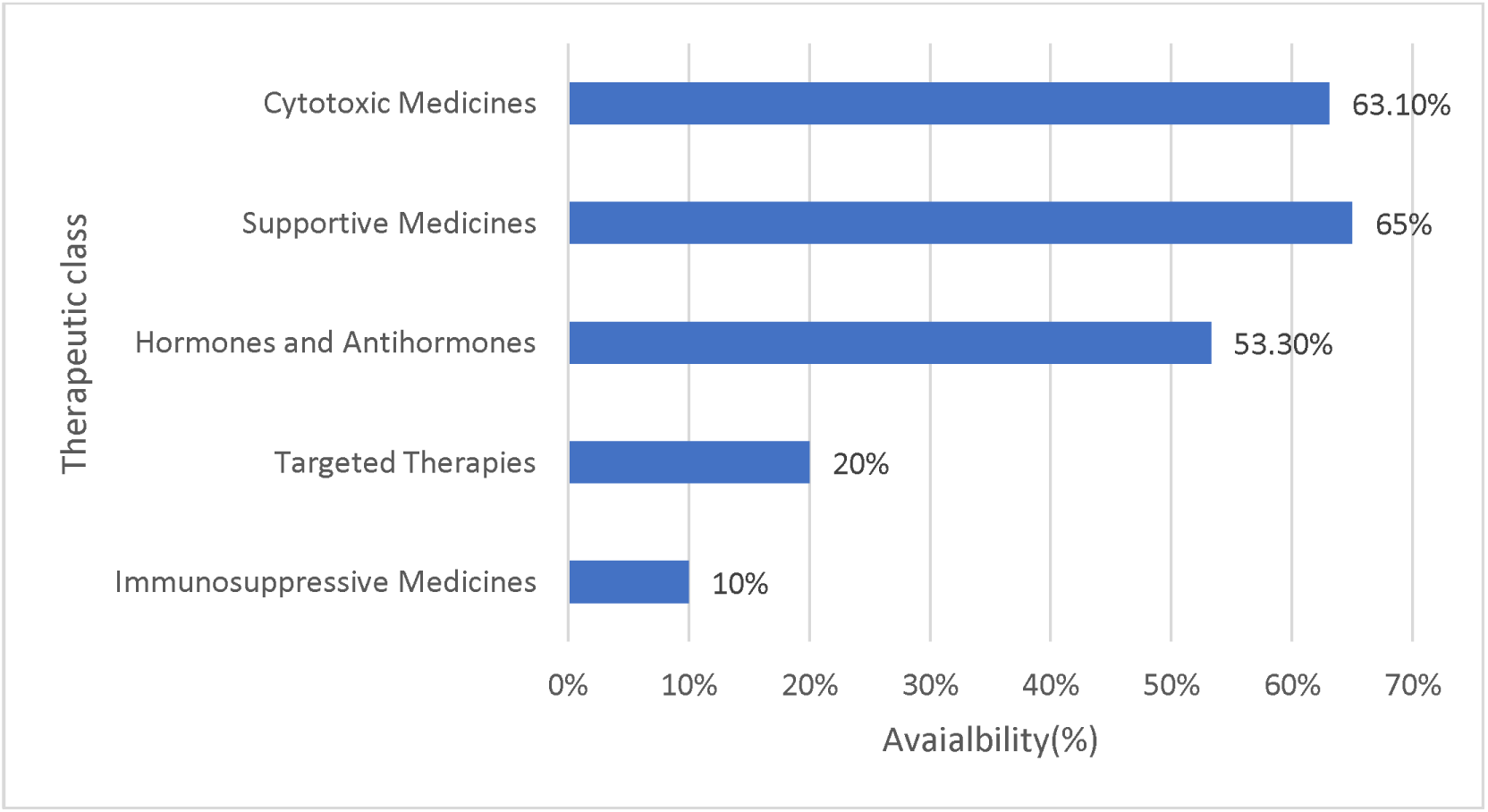
Availability of cancer medicines by therapeutic class.

Among immunosuppressives, only ciclosporin 25 mg and tacrolimus 1 mg were available, and only at private facility 1 (20%). For targeted therapies, only bortezomib (80%) and rituximab (60%) demonstrated moderate stocking. Erlotinib, gefitinib, osimertinib, ibrutinib, nilotinib, and all-trans retinoic acid were completely unavailable across all facilities surveyed. Imatinib was available through donation only at public facility 1, while at private facility 1 it was stocked as a generic (Mytinib). Among hormonal therapies, Letrozole and tamoxifen were available in 80% of facilities. Bicalutamide showed 40% availability. Supportive medicines were among the most available, with filgrastim and zoledronic acid each available in 80% of facilities, and allopurinol in 60%. For cytotoxic medicines, cisplatin, cyclophosphamide, doxorubicin (conventional), paclitaxel, and oxaliplatin were available across all five facilities (100%), but several others showed limited availability (20%), and L-asparaginase and mercaptopurine were entirely absent. (Table 1)

**Table 1:**
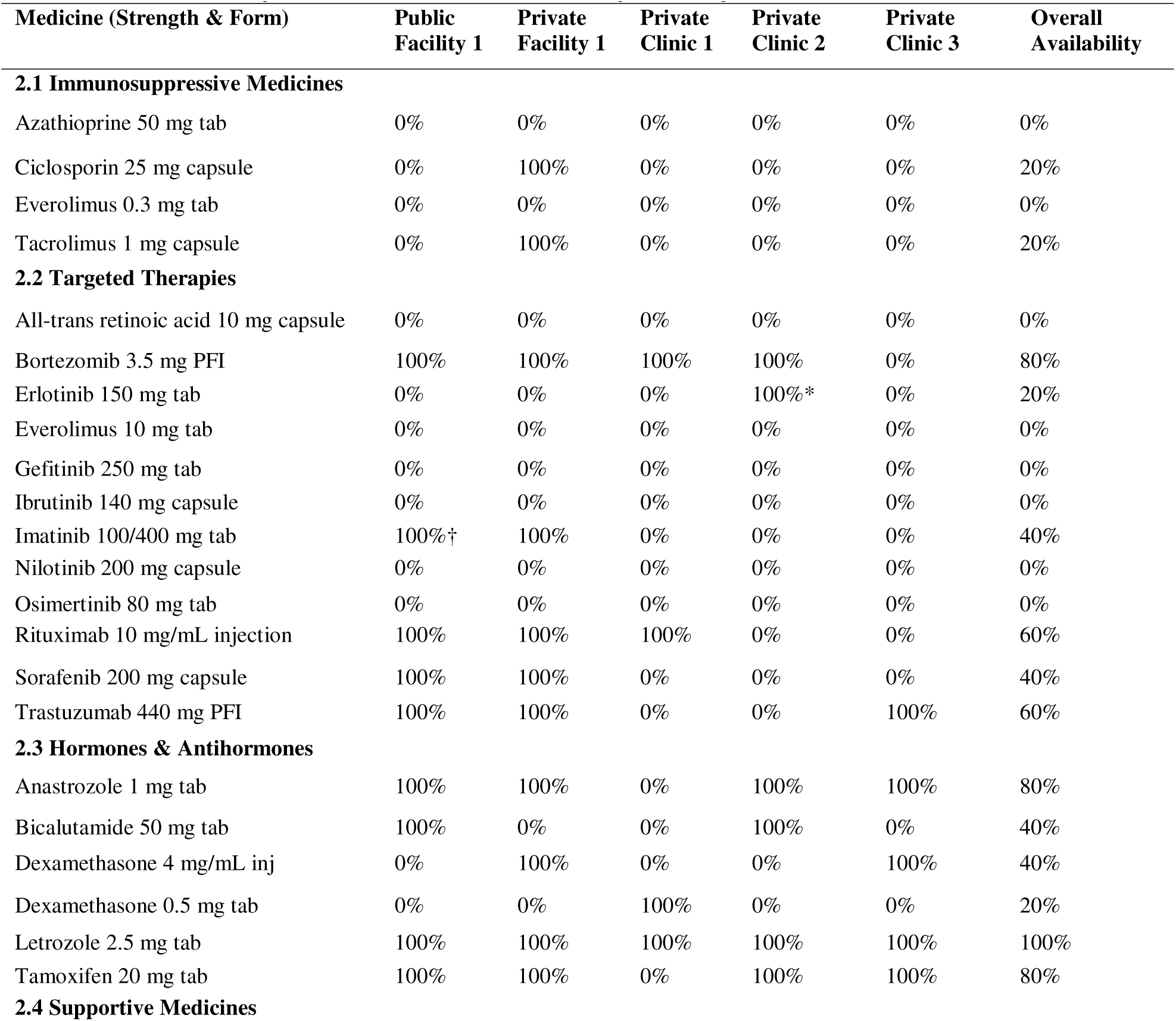
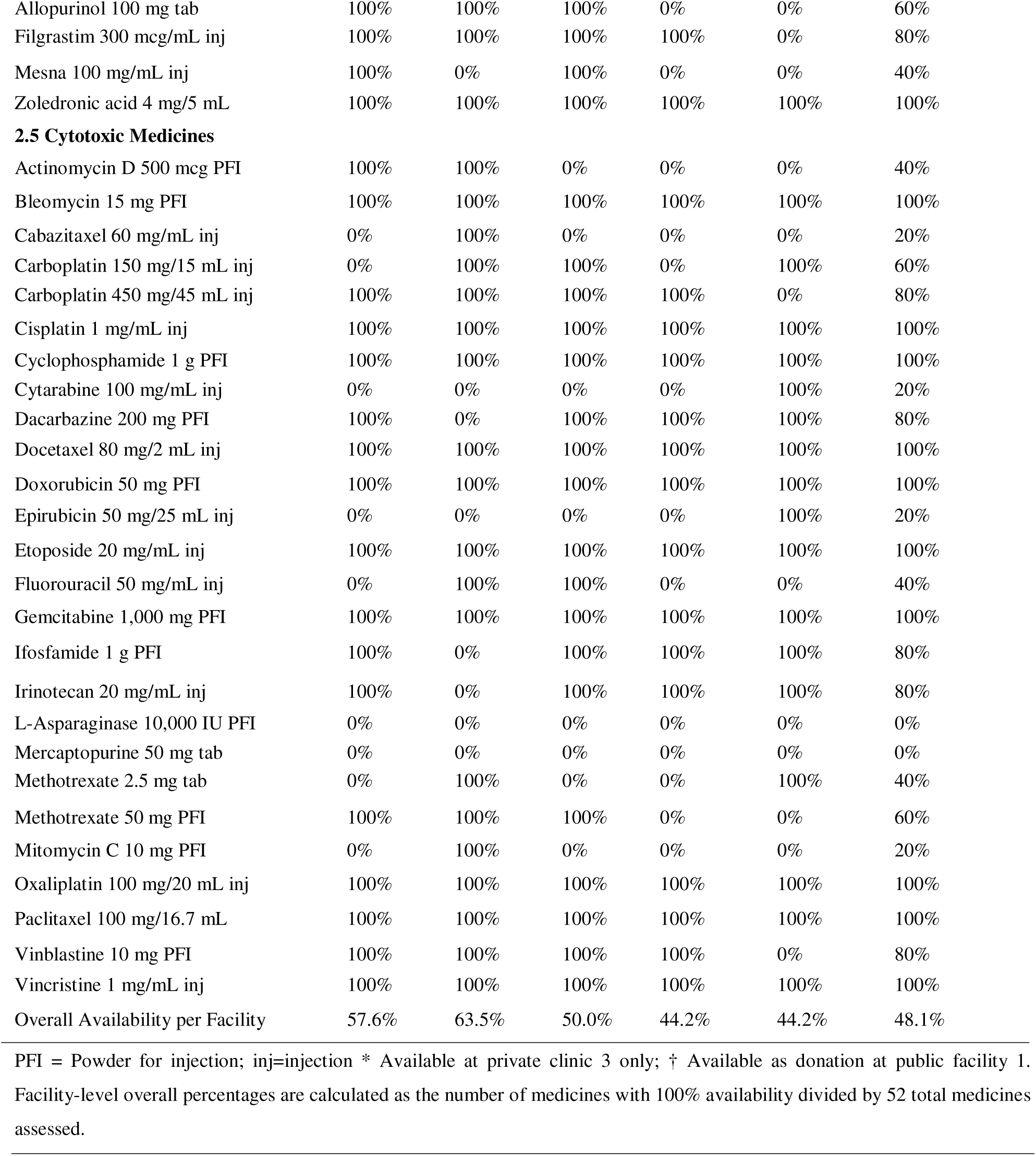
Availability of Anticancer Medicines by Facility and Overall.

### 3.2 Originator Brand versus Generic Availability

Generic (low-priced generic [LPG]) medicines were substantially more available than originator brand equivalents across all therapeutic classes. Overall, LPGs demonstrated an average availability of 41.8% compared to 7.3% for originator brands, a more than fivefold difference.

LPG availability was highest for cytotoxic medicines (63.1%) and supportive medicines (65%), while originator availability was negligible or zero in most classes. (Table 2)

**Table 2:** Originator Brand vs. Generic Medicine Availability by Therapeutic Class.

| Therapeutic Group | No. of Medicines | Originator Avg. Availability (%) | LPG Avg. Availability (%) |
| --- | --- | --- | --- |
| 2.1 Immunosuppressive Medicines | 4 | 0% | 10.0% |
| 2.2 Targeted Therapies | 12 | 13.3% | 20.0% |
| 2.3 Hormones & Antihormones | 6 | 16.7% | 53.3% |
| 2.4 Supportive Medicines | 4 | 10.0% | 65.0% |
| 2.5 Cytotoxic Medicines | 26 | 0% | 63.1% |
| Overall Average | 52 | 7.3% | 41.8% |

### 3.3 Cancer Medicine Prices and Inter-Facility Variation

Immunosuppressive medicines where available were uniformly low-cost (unit prices < 0.1–0.55 days’ wages). Targeted therapies imposed the heaviest financial burden. Rituximab demonstrated extreme price variation across facilities, ranging from KES 54,000 (USD 417) to KES 142,000 (USD 1,096.5) per 500 mg vial equivalent, requiring 73.7 to 193.9 days’ wages per single dose. Trastuzumab (440 mg vial) ranged from KES 40,000 (USD 308.9) to KES 66,000 (USD 509.7) (54.6–90.1 days’ wages), and bevacizumab (400 mg vial) from KES 50,500 (USD 390) to KES 63,000 (USD 486.9) (69.0–86.0 days’ wages). Notably, the public facility offered bortezomib at KES 22,000 (USD 169.9) per vial, nearly double the price at one private clinic (KES 11,900; USD 91.9).

Hormonal therapies were relatively affordable overall, though generic anastrozole showed up to approximately 26-fold variation (KES 100–2,557; USD 0.8–19.7). The anastrozole originator brand at public facility 1 (KES 5,200) was separately reported. Comparing originator to generic yields a 52-fold headline range (KES 100–5,200; USD 0.8–40.2). For letrozole, generic prices ranged from KES 70–2,557 (USD 0.5–19.7) and the originator at public facility 1 reaching KES 7,650 (USD 59.1). Octreotide (30 mg vial), available at private facility 1, was the single most expensive supportive medicine at KES 60,000 (USD 463.3; 81.9 days’ wages).

Standard cytotoxic agents showed moderate affordability but pronounced inter-facility variation, reaching 3.6-fold for oxaliplatin (KES 2,475–9,000; USD 19–69) and more than 6-fold for docetaxel (KES 2,500–16,000; USD 19–123). Carboplatin (450 mg) ranged from KES 5,650-12,000; (USD 43.6-92.7), Docetaxel (80 mg) from KES 2,500-16,000 (USD 19.3-123.6). (Table 3)

**Table 3:** Prices of Selected Cancer Medicines by Facility.

| Medicine (Strength & Form) | Public Facility 1) KES (USD) | Private Facility 1) KES (USD) | Private Clinic 1) KES (USD) | Private Clinic 2(KES (USD) | Private Clinic 3 KES (USD) | Price Range KES (USD) | Days' Wages (Min–Max) |
| --- | --- | --- | --- | --- | --- | --- | --- |
| **Immunosuppressives** |  |  |  |  |  |  |  |
| Azathioprine 50 mg tab (unit) | N/A | 60 (0.5) | N/A | N/A | N/A | 60 (0.5) | 0.08 |
| Ciclosporin 25 mg capsule (unit) | N/A | 400 (3.1) | N/A | N/A | N/A | 400 (3.1) | 0.55 |
| Tacrolimus 1 mg capsule (unit) | N/A | 100 (0.8) | N/A | N/A | N/A | 100 (0.8) | 0.14 |
| **Targeted Therapies** |  |  |  |  |  |  |  |
| Bortezomib 3.5 mg vial | 22,000 (169.9) | 13,800 (106.6) | 11,900 (91.9) | N/A | N/A | 11,900–22,000 (91.9–169.9) | 16.2–30.0 |
| Erlotinib 150 mg tab | N/A | N/A | N/A | 13,736 (106.1) | N/A | 13,736 (106.1) | 18.8 |
| Imatinib tab (generic, unit) | Donation | 127.5 (1.0) | N/A | N/A | N/A | 127.5 (1.0) | 0.17 |
| Rituximab 500 mg vial equiv. | 60,000 (463.3) | 54,000 (417.0) | 63,000 (486.9) | N/A | N/A | 54,000–142,000* (417–1,096.5) | 73.7–193.9 |
| Sorafenib 200 mg capsule | 7,500 (57.9) | 4,000 (30.9) | N/A | N/A | N/A | 4,000–7,500 (30.9–57.9) | 5.5–10.2 |
| Trastuzumab 440 mg vial | 50,000 (386.1) | 40,000 (308.9) | N/A | 45,000 (347.5) | 66,000 (509.7) | 40,000–66,000 (308.9–509.7) | 54.6–90.1 |
| Bevacizumab 400 mg vial | N/A | N/A | 63,000 (486.9) | 50,500 (390.0) | N/A | 50,500–63,000 (390.0–486.9) | 69.0–86.0 |
| **Hormones & Antihormones** |  |  |  |  |  |  |  |
| Anastrozole 1 mg tab (generic) | — | 100 (0.8) | N/A | 2,500 (19.3) | 2,557 (19.7) | 100–2,557 (0.8–19.7) | 0.14–3.5 |
| Anastrozole 1 mg tab (originator) | 5,200 (40.2) | — | N/A | — | — | — | — |
| Bicalutamide 50 mg tab | 12,000 (92.7) | N/A | N/A | 3,000 (23.2) | 8,250 (63.7) | 3,000–12,000 (23.2–92.7) | 4.1–16.4 |
| Letrozole 2.5 mg tab (generic) | — | 70 (0.5) | 2,500 (19.3) | 2,500 (19.3) | 2,557 (19.7) | 70–2,557 (0.5–19.7) | 0.10–3.5 |
| Letrozole 2.5 mg tab (originator) | 7,650 (59.1) | — | — | — | — | — | — |
| Tamoxifen 20 mg tab | 1,800<br>(13.9) | 15 (0.1) | N/A | 3,000<br>(23.2) | 1,443<br>(11.1) | 15–3,000<br>(0.1–23.2) | 0.02–4.1 |
| Abiraterone 250 mg tab<br>(120s) | 50,000<br>(386.1) | N/A | 60,000<br>(463.3) | N/A | N/A | 50,000–60,000<br>(386.1–463.3) | 68.3–81.9 |
| Enzalutamide 80 mg tab<br>(14s) | 20,000<br>(154.4) | N/A | N/A | N/A | N/A | 20,000<br>(154.4) | 27.3 |
| Supportive Medicines |  |  |  |  |  |  |  |
| Allopurinol 100 mg tab | N/A | 8–50<br>(0.1–0.4) | N/A | 600 (4.6) | N/A | 8–600<br>(0.1–4.6) | 0.01–0.82 |
| Filgrastim 300 mcg vial | 3,000<br>(23.2) | 4,400<br>(34.0) | 2,500<br>(19.3) | 2,500<br>(19.3) | N/A | 2,500–4,400<br>(19.3–34.0) | 3.4–6.0 |
| Zoledronic acid 4 mg vial | 4,150<br>(32.0) | 4,000<br>(30.9) | 1,800<br>(13.9) | 1,800<br>(13.9) | 1,237<br>(9.6) | 1,237–4,150<br>(9.6–32.0) | 1.7–5.7 |
| Octreotide 30 mg vial | N/A | 60,000<br>(463.3) | 51,000<br>(393.8) | N/A | N/A | 51,000–60,000<br>(393.8–463.3) | 69.6–81.9 |
| **Selected Cytotoxic**<br>**Medicines** |  |  |  |  |  |  |  |
| Bleomycin 15 mg vial | 3,500<br>(27.0) | 3,800<br>(29.3) | 2,500<br>(19.3) | 2,500<br>(19.3) | 2,310<br>(17.8) | 2,310–4,960<br>(17.8–38.3) | 3.2–6.8 |
| Carboplatin 450 mg vial | 6,000<br>(46.3) | 12,000<br>(92.7) | 5,650<br>(43.6) | 7,920<br>(61.2) | N/A | 5,650–12,000<br>(43.6–92.7) | 7.7–16.4 |
| Cisplatin 50 mg vial | 2,700<br>(20.8) | 2,000<br>(15.4) | 3,000<br>(23.2) | 1,200<br>(9.3) | 1,897<br>(14.6) | 1,200–3,000<br>(9.3–23.2) | 1.6–4.1 |
| Cyclophosphamide 1 g vial | 1,250 (9.7) | 1,000<br>(7.7) | 1,300<br>(10.0) | 560 (4.3) | 701<br>(5.4) | 560–1,300<br>(4.3–10.0) | 0.8–1.8 |
| Docetaxel 80 mg vial | N/A | 16,000<br>(123.6) | 2,500<br>(19.3) | 2,500<br>(19.3) | 8,250<br>(63.7) | 2,500–16,000<br>(19.3–123.6) | 3.4–21.8 |
| Doxorubicin 50 mg vial<br>(conventional) | 1,250 (9.7) | 1,700<br>(13.1) | 1,500<br>(11.6) | 1,200<br>(9.3) | N/A | 1,200–1,700<br>(9.3–13.1) | 1.6–2.3 |
| Doxorubicin liposomal 50<br>mg | N/A | N/A | N/A | N/A | 36,093<br>(278.7) | 36,093 (278.7) | 49.3 |
| Gemcitabine 1,000 mg vial | 5,500<br>(42.5) | 6,000<br>(46.3) | 5,500<br>(42.5) | 4,800<br>(37.1) | 1,815<br>(14.0) | 1,815–6,000<br>(14.0–46.3) | 2.5–8.2 |
| Irinotecan 20 mg/mL (5 mL<br>vial) | 6,450<br>(49.8) | N/A | 6,000<br>(46.3) | N/A | 3,465<br>(26.8) | 3,465–6,450<br>(26.8–49.8) | 4.7–8.8 |
| Oxaliplatin 100 mg vial | 7,200<br>(55.6) | 9,000<br>(69.5) | 4,000<br>(30.9) | 4,200<br>(32.4) | 2,475<br>(19.1) | 2,475–9,000<br>(19.1–69.5) | 3.4–12.3 |
| Paclitaxel 100 mg vial | 3,500<br>(27.0) | 3,000<br>(23.2) | 4,500<br>(34.7) | 5,880<br>(45.4) | 2,887<br>(22.3) | 2,887–5,880<br>(22.3–45.4) | 3.9–8.0 |
| Vincristine 1 mg vial | 900 (7.0) | 1,000 (7.7) | 450 (3.5) | 429 (3.3) | 2,887 (22.3) | 429–2,887 (3.3–22.3) | 0.6–3.9 |

Rituximab originator (Mabthera) at private facility 1 = KES 142,000 (USD 1,096.5; 193.9 days' wages). †private clinic 2/3 Trastuzumab price varies across sub-facilities (KES 45,000–66,000). Anastrozole and Letrozole: Generic and originator prices shown separately where both exist.

### 3.4 Median Patient Prices, and Median Price Ratios (MPR) for Selected Cancer Medicines

Prices were further benchmarked against UNICEF supply division indicative procurement prices for quality-assured generics. MPR was calculated as the ratio of the local median patient selling price (converted to USD) to the corresponding UNICEF indicative price. For the medicines with confirmed UNICEF matches, lowest local prices in Kisumu County were competitive for several essential agents. Cyclophosphamide achieved a median MPR of 0.53, vincristine 0.38, filgrastim 0.45, and allopurinol 0.30. In contrast, platinum compounds showed higher ratios, with cisplatin recording a median MPR of 1.42–2.64 and carboplatin 1.90–4.07 depending on vial size equivalence. Substantial inter-facility price variation resulted in MPRs at the highest-cost facilities often exceeding UNICEF benchmarks by two-to six-fold. Table 4

**Table 4:** Median Patient Prices, and Median Price Ratios (MPR) for Selected Cancer Medicines.

| Medicine (Strength & Form) | Local Median Price (USD) | Local Price Range (USD) | UNICEF Indicative Price (USD) | MPR (Median) | MPR Range (Min–Max) |
| --- | --- | --- | --- | --- | --- |
| Cisplatin 1 mg/mL (10 mL vial) | 18.5 | 9.3 – 23.2 | 7 | 2.64 | 1.33 – 3.31 |
| Cisplatin 1 mg/mL (50 mL vial) | 18.5 | 9.3 – 23.2 | 13 | 1.42 | 0.72 – 1.78 |
| Carboplatin 10 mg/mL (40 mL vial) | 61 | 43.6 – 92.7 | 32.04 | 1.9 | 1.36 – 2.89 |
| Cyclophosphamide 1 g powder for injection | 7.7 | 4.3 – 10.0 | 14.6 | 0.53 | 0.29 – 0.68 |
| Vincristine 1 mg/mL injection | 7 | 3.3 – 22.3 | 18.4 | 0.38 | 0.18 – 1.21 |
| Filgrastim (various strengths) | 23.2 | 19.3 – 34.0 | 45.73 – 80.76 | 0.45 | 0.24 – 0.74 |
| Allopurinol 100 mg tablet | 0.3 | 0.1 – 4.6 | 1 | 0.3 | 0.10 – 4.60 |

\*UNICEF/International Reference: Based on recent UNICEF Supply Division/Global Platform tender benchmarks for quality-assured generics and comparable LMIC pooled procurement (2023–2025 data). Ranges reflect typical awarded tender prices. MPR < 1.0 = below reference; MPR 1.0–2.0 = moderate; >2.0 = high mark-up. Targeted therapies (rituximab, trastuzumab) and many orals excluded due to limited comparable UNICEF adult references. Notes: MPR = local median patient price ÷ UNICEF indicative price. Local prices are patient selling prices recorded across the five facilities in October 2025

### 3.5 Chemotherapy Regimen Costs for One Cycle

Regimen costs were calculated using BSA = 1.7 m² and lowest available generic prices at each facility. Full vials were priced at listed patient prices with no pro-rating. Where the calculated dose exceeded a single vial, the minimum number of whole vials required was used. The breast cancer AC regimen cost ranged from KES 3,560-5800 (USD 27-45; 4.9-7.9 days’ wages), a 1.6-fold range. Weekly cisplatin for cervical cancer was the most affordable, ranging from KES 2,400–6,000 (USD 18.5–46.3; 3.3–8.2 days’ wages). Prostate cancer docetaxel ranged from KES 5,000–32,000 (USD 38.6–247.1; 6.8–43.7 days’ wages). Oesophageal cancer cisplatin+5-FU (q3w) cost KES 8,760–15,800 (USD 67.6–122.0; 12.0–21.6 days’ wages). Colorectal FOLFOX (q2w) ranged from KES 7,350–18,500 (USD 56.8–142.9; 10.0–25.3 days’ wages), excluding leucovorin. (Table 5)

**Table 5:** Cost Per One Chemotherapy Cycle Per Facility.

| Cancer Type & Regimen (1 cycle) | Public Facility) KES (USD) | Private Facility.) KES (USD) | Private Clinic 1) KES (USD) | Private Clinic 2 KES (USD) | Private Clinic 3 KES (USD) | Price Range KES (USD) | Days' Wages (Min–Max) |
| --- | --- | --- | --- | --- | --- | --- | --- |
| Breast – AC (q3w)<br>Doxorubicin 60 mg/m<sup>2</sup> +<br>Cyclophosphamide 600 mg/m<sup>2</sup> | 5,000 (38.6) | 4,700 (36.3) | 3,560 (27.5) | 5,800 (44.8) | N/Aa | KES 3,560–5,800 (27–45) | 4.9–7.9 |
| Cervical – Weekly<br>Cisplatin 40 mg/m<sup>2</sup> (68 mg/dose) | 5,400 (41.7) | 4,000 (30.9) | 6,000 (46.3) | 2,400 (18.5) | 3,794 (29.3) | 2,400–6,000 (18.5–46.3) | 3.3–8.2 |
| Prostate – Docetaxel (q3w) † 75 mg/m<sup>2</sup> (127.5 mg → 2×80 mg vials) | 5000 (38.6) | 32,000 (247.1) | 5,000 (38.6) | 5,000 (38.6) | 16,500 (127.4) | 5,000–32,000 (38.6–247.1) | 6.8–43.7 |
| Oesophageal – Cisplatin + 5-FU (q3w) Cisplatin 75 mg/m<sup>2</sup> + 5-FU 1,000 mg/m<sup>2</sup> D1–4 | 15,800 (122.0) | 14,200 (109.7) | 8,760 (67.6) | 12,500 (96.5) | 9,675§ (74.7) | 8,760–15,800 (67.6–122.0) | 12.0–21.6 |
| Colorectal – FOLFOX (q2w) ¶ Oxaliplatin 85 mg/m<sup>2</sup> + 5-FU bolus + infusion | 14,475 (111.8) | 18,500 (142.9) | 7,350 (56.8) | 9,200 (71.0) | 8,450§ (65.3) | 7,350–18,500 (56.8–142.9) | 10.0–25.3 |

¶ Leucovorin (400 mg/m<sup>2</sup>) cost not included (estimated additional KES 2,000–14,000 per cycle).

### 3.6 Full-Course Affordability Analysis

The affordability burden increased substantially when complete treatment courses were considered. Depending on the regimen and facility, full-course treatment required between 19.6 and 437 days’ wages. Breast cancer AC and weekly cisplatin for cervical cancer required 19.6–47.4 and 19.8–49.2 days’ wages, respectively. In contrast, prostate cancer docetaxel required up to 437 days’ wages, while colorectal FOLFOX required 80.0–303.6 days’ wages and oesophageal CF required 36.0–129.6 days’ wages for a complete course of treatment. (Table 6)

**Table 6:**
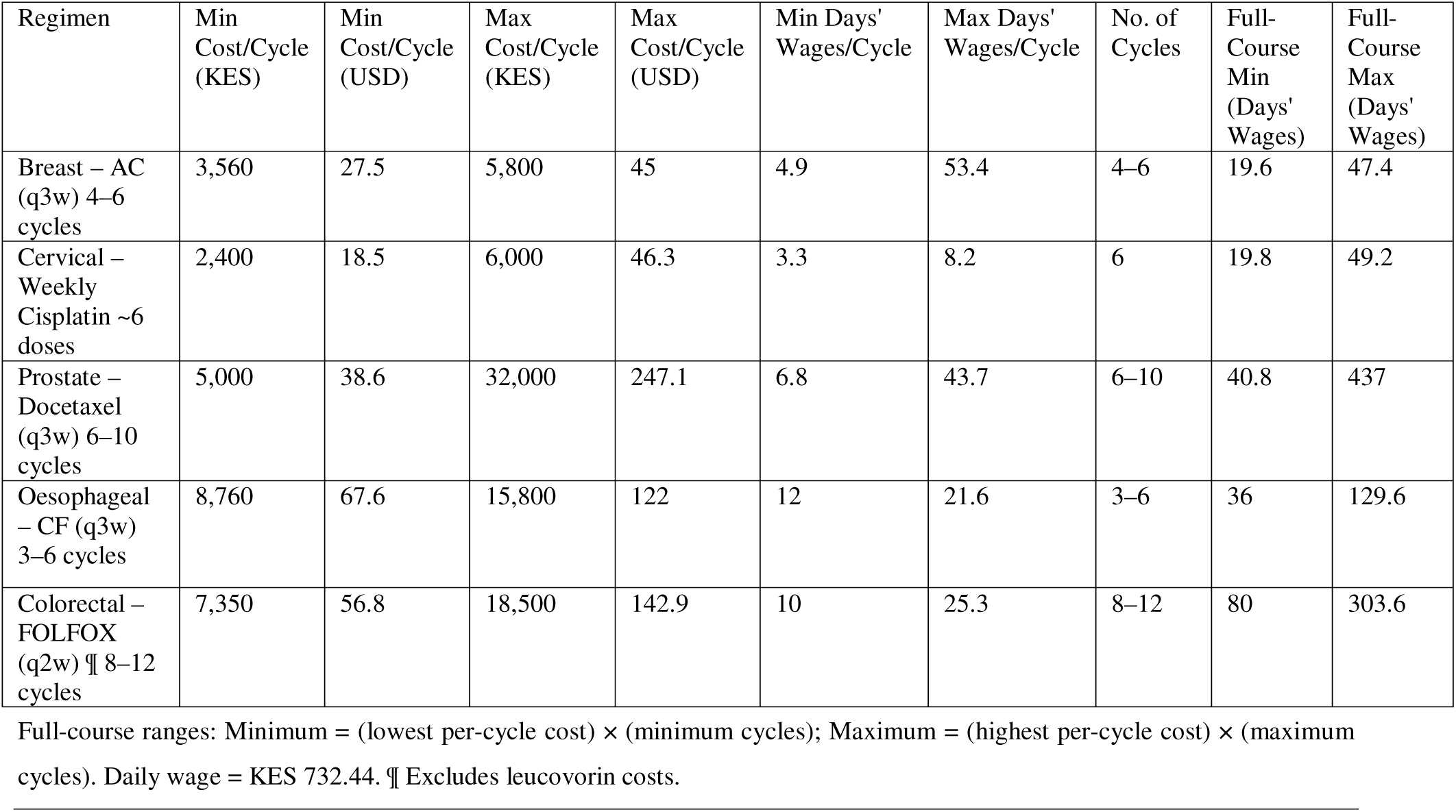
Full-Course Treatment Cost Ranges by Regimen (Days’ Wages).

| Regimen | Min Cost/Cycle (KES) | Min Cost/Cycle (USD) | Max Cost/Cycle (KES) | Max Cost/Cycle (USD) | Min Days' Wages/Cycle | Max Days' Wages/Cycle | No. of Cycles | Full-Course Min (Days' Wages) | Full-Course Max (Days' Wages) |
| --- | --- | --- | --- | --- | --- | --- | --- | --- | --- |
| Breast – AC (q3w) 4–6 cycles | 3,560 | 27.5 | 5,800 | 45 | 4.9 | 53.4 | 4–6 | 19.6 | 47.4 |
| Cervical – Weekly Cisplatin ~6 doses | 2,400 | 18.5 | 6,000 | 46.3 | 3.3 | 8.2 | 6 | 19.8 | 49.2 |
| Prostate – Docetaxel (q3w) 6–10 cycles | 5,000 | 38.6 | 32,000 | 247.1 | 6.8 | 43.7 | 6–10 | 40.8 | 437 |
| Oesophageal – CF (q3w) 3–6 cycles | 8,760 | 67.6 | 15,800 | 122 | 12 | 21.6 | 3–6 | 36 | 129.6 |
| Colorectal – FOLFOX (q2w) ¶ 8–12 cycles | 7,350 | 56.8 | 18,500 | 142.9 | 10 | 25.3 | 8–12 | 80 | 303.6 |

Full-course ranges: Minimum = (lowest per-cycle cost) × (minimum cycles); Maximum = (highest per-cycle cost) × (maximum cycles). Daily wage = KES 732.44. ¶ Excludes leucovorin costs.

### 3.7 SHA Benefit Package Adequacy Analysis

We accessed full-course regimen costs as a percentage of the KSh 550,000 ceiling, using both best-case (lowest facility price × minimum cycles) and worst-case (highest facility price × maximum cycles) scenarios.

Standard cytotoxic regimens for breast, cervical and oesophageal cancers were generally adequately covered within the annual benefit ceiling. Breast cancer AC (4–6 cycles) costs KES 14,347–34,693, consuming only 2.6–6.3% of the annual cap. Weekly cisplatin for cervical cancer (6 doses) costs KES 14,494–36,026 (2.6–6.5% of cap). Oesophageal cisplatin-5FU (3–6 cycles) costs KES 26,368–94,892 (4.8–17.3% of cap).

However, targeted therapies imposed the greatest pressure on the SHA benefit package. Prostate cancer docetaxel (6–10 cycles) costs KES 29,874–319,918 (5.4–58.2% of cap), creating meaningful uncertainty at high prices. Colorectal FOLFOX (8–12 cycles, excluding leucovorin) costs KES 58,595–222,235 (10.7–40.4% of cap), leaving limited headroom once diagnostics, supportive medicines, and leucovorin are added. The SHA ceiling is critically stressed or exceeded when targeted therapies are required. For HER2-positive breast cancer, trastuzumab alone (17–18 cycles × KES 40,000–66,000/vial) costs KES 680,000–1,188,000 equivalent to 23.6% to 116% above the SHA annual cap before adding the cost of AC chemotherapy. For recurrent or metastatic cervical cancer, six doses of bevacizumab cost KES 303,000–378,000 (55–69% of the SHA cap), effectively exhausting coverage for concurrent diagnostic and supportive costs. For metastatic prostate cancer, four months of abiraterone alone costs KES 200,000–240,000 (36–44% of SHA cap), and continuous monthly dosing is required indefinitely for responders. (Table 7)

**Table 7:** Full-Course Regimen Costs vs. SHA KSh 550,000 Annual Benefit Package.

| Cancer / Regimen | Projected Min Full-Course (KES) | Projected Max Full-Course (KES) | Min as % of SHA Cap | Max as % of SHA Cap | Adequacy (Chemotherapy Only) |
| --- | --- | --- | --- | --- | --- |
| Breast – AC (q3w), 4–6 cycles | 14,347 | 34,693 | 2.60% | 6.30% | Adequate |
| Cervical – Weekly Cisplatin, ~6 doses | 14,494 | 36,026 | 2.60% | 6.50% | Adequate |
| Oesophageal – CF (q3w), 3–6 cycles | 26,368 | 94,892 | 4.80% | 17.30% | Likely adequate |
| Prostate – Docetaxel (q3w), 6–10 cycles | 29,874 | 319,918 | 5.40% | 58.20% | Borderline (variable by facility) |
| Colorectal – FOLFOX (q2w), 8–12 cycles¶ | 58,595 | 222,235 | 10.70% | 40.40% | Risky at high-price facilities |
| HER2+ Breast – Trastuzumab adjuvant (17–18 cycles) | 680,000 | 1,188,000 | 123.60% | 216.00% | Exceeds SHA cap (chemo not yet added) |
| Recurrent Cervical – Bevacizumab (6 doses) | 303,000 | 378,000 | 55.10% | 68.70% | Severely strained /cap nearly exhausted by drug alone |

¶FOLFOX excludes leucovorin; adding leucovorin adds up to KES 14,000/cycle. Trastuzumab and Bevacizumab rows assess targeted therapy alone; backbone chemotherapy costs are additive. SHA package assessed against chemotherapy drug costs only.

### 3.8 Sensitivity Analysis of Regimen Cost Adequacy under the Revised SHA KES 800,000 Cap

A presidential directive raised the SHA oncology benefit cap from Kenya shillings (KES) 550,000 to KES 800,000 gazetted under Legal Notice No. 78 of 2026 (signed 30 April 2026, gazetted 8 May 2026) and took effect immediately. Because this change occurred after our facility-level price survey (October 2025), we re-applied the regimen costs reported against the new KES 800,000 ceiling using the same methodology, to assess whether the increase resolves the affordability gaps identified in the original analysis.

Applying the revised SHA oncology benefit ceiling of KES 800,000 improved coverage across all treatment regimens. Standard cytotoxic regimens for breast, cervical, oesophageal and colorectal cancers were adequately covered well under 30% of the annual ceiling even at maximum observed facility prices. Nevertheless, affordability challenges persisted for high-cost targeted therapies. Trastuzumab remained above the annual benefit ceiling at the highest observed facility prices, while annualized abiraterone therapy consumed 75–90% of the revised package. Bevacizumab became affordable as a single medicine under the new ceiling but continued to leave limited financial capacity for concurrent diagnostics, supportive medicines and other components of comprehensive cancer care incurred in the same benefit year. (Table 8)

**Table 8:** Full-Course Regimen Costs vs. Revised SHA KSh 800,000 Annual Benefit Package.

| Cancer / Regimen | Projected Min Full-Course (KES) | Projected Max Full-Course (KES) | Min as % of New SHA Cap | Max as % of New SHA Cap | Adequacy (Chemotherapy Only) |
| --- | --- | --- | --- | --- | --- |
| Breast – AC (q3w), 4–6 cycles | 14,347 | 34,693 | 1.79% | 4.34% | Adequate |
| Cervical – Weekly Cisplatin, ~6 doses | 14,494 | 36,026 | 1.81% | 4.50% | Adequate |
| Oesophageal – CF (q3w), 3–6 cycles | 26,368 | 94,892 | 3.30% | 11.86% | Adequate |
| Prostate – Docetaxel (q3w), 6–10 cycles | 29,874 | 319,918 | 3.73% | 39.99% | Borderline (variable by facility) |
| Colorectal – FOLFOX (q2w), 8–12 cycles¶ | 58,595 | 222,235 | 7.32% | 27.78% | Adequate to moderate risk at high-price facilities |
| HER2+ Breast – Trastuzumab adjuvant (17–18 cycles) | 680,000 | 1,188,000 | 85.00% | 148.50% | Improved, but still exceeds cap at high-price facilities (chemo not yet added) |
| Recurrent Cervical – Bevacizumab (6 doses) | 303,000 | 378,000 | 37.88% | 47.25% | Adequate for drug alone; limits headroom for diagnostics/supportive care |
| Metastatic Prostate –<br>Abiraterone (continuous), 12<br>months annualised* | 600,000 | 720,000 | 75.00% | 90.00% | Severely strained;<br>single continuous agent<br>alone approaches full<br>cap |

¶FOLFOX excludes leucovorin; adding leucovorin adds up to KES 14,000/cycle. \*Abiraterone row is not a discrete full-course regimen from the original per-cycle costing framework; it is extrapolated by annualising the per-month price data reported in over 12 months of continuous therapy, to illustrate exposure from a single continuous agent under an annual ceiling. Trastuzumab and Bevacizumab rows assess targeted therapy alone; backbone chemotherapy costs are additive. SHA package assessed against chemotherapy/targeted-therapy drug costs only; diagnostics, radiotherapy, and supportive medicines draw on the same undifferentiated annual cap.

## 4. DISCUSSION

### 4.1 Availability of Cancer Medicines

This study found that the overall availability of essential cancer medicines in Kisumu County was 48.1%, below the WHO target of at least 80%. This finding indicates that access to cancer medicines remains suboptimal despite efforts to strengthen cancer services in Kenya. The observed availability is better than comparable findings from LMIC settings. Alemu and Hailemariam (2022) reported availability of approximately 20% for anticancer medicines in Addis Ababa, Ethiopia (16). Higgins et al. (2026), surveying Cameroon, Ethiopia, Kenya, and Malawi, documented chemotherapy stockouts in public hospitals in three of four countries (7). Likewise, Kizub et al. (2022) found that availability of WHO-listed essential cancer medicines in Kenya, Rwanda, and Uganda fell below levels needed for uninterrupted care (8).

Availability differed between therapeutic classes. Conventional cytotoxic medicines and supportive medicines were relatively well stocked, whereas targeted therapies and immunosuppressive medicines showed markedly lower availability. This pattern is consistent with previous findings that public procurement systems preferentially stocked older cytotoxic medicines because they are included in national essential medicines lists, available as generics and less expensive than targeted therapies (8, 17). Similarly, Cherny et al. (2017) reported that access to innovative cancer medicines remains limited in LMICs because of high acquisition costs and delayed incorporation into national reimbursement programs (18). The result is a two-tier availability landscape. Patients with common solid tumors may access basic cytotoxic regimens. Those requiring targeted therapies, immunotherapies, or biologic agents face near-complete unavailability. Patients redirected from public facilities during stockouts encounter private clinics offering identical protocols at higher costs and may exhaust their annual SHA oncology benefit within a fraction of the prescribed treatment course.

The absence of several medicines, including everolimus, L-asparaginase and mercaptopurine, suggests persistent weaknesses in procurement planning and supply chain management. Such shortages may result from inadequate forecasting, irregular supplier availability and limited procurement budgets. These deficiencies have important clinical implications because interruptions in medicine availability can delay treatment initiation, necessitate regimen modification and ultimately compromise patient outcomes. Overall, ensuring sustainable financing for essential oncology medicines should remain a priority for improving cancer care in Kenya.

A key finding of this study was that generic medicines were more available than originator brands across all therapeutic classes. Procurement practices increasingly favour lower-priced generic products to maximise medicine coverage within constrained healthcare budgets. Previous findings indicate that generic medicines constitute the majority of available oncology products because they offer considerable cost savings while maintaining therapeutic equivalence (7,8). The predominance of generic medicines is encouraging from both clinical and economic perspectives. WHO recommends the use of quality-assured generics as an effective strategy for improving medicine accessibility and reducing pharmaceutical expenditure (19). However, the low availability of originator products also reflects the limited market penetration of newer patented medicines, particularly targeted therapies. Many innovative oncology medicines remain protected by intellectual property rights, have limited biosimilar competition and are sold at prices that exceed the purchasing capacity of many public and private healthcare providers in Kenya (20). Thus, complementary interventions aimed at strengthening financing mechanisms for high-cost oncology medicines would substantially improve equitable access to essential cancer treatment in Kenya and similar settings.

### 4.2 Price Variability and Structural Drivers

This study demonstrated substantial inter-facility variation in the prices of cancer medicines, with the magnitude of variation differing considerably across therapeutic classes. Although conventional cytotoxic medicines were generally less expensive than targeted therapies, considerable price differences existed even for identical medicines and strengths, suggesting inconsistent procurement practices and medicine pricing across facilities. In a multi-country price analysis of childhood cancer chemotherapy prices, three-to sixfold price variation for commonly used anticancer medicines were documented across facilities despite identical formulations (21). Similarly, Ocran Mattila et al. (2021), in a systematic review of anti-cancer medicine pricing, documented intra-country price differentials of up to 700% across LMIC facilities (22). The authors attributed these disparities to fragmented procurement systems, differences in supplier contracts, variable mark-ups, and the absence of effective price regulation. Thus, formal price-regulation frameworks alone, without complementary retail mark-up controls and pooled procurement, may not be sufficient to eliminate price differences.

The greatest price disparities were observed for targeted therapies, particularly rituximab, trastuzumab and bevacizumab, which required several months’ wages for a single dose. These findings reflect the continuing challenge of accessing biological medicines in many LMICs. Unlike conventional cytotoxic medicines, biologics are expensive to manufacture, frequently remain under patent protection, have limited biosimilar competition and are associated with complex procurement and cold-chain requirements (20). Consequently, even where these medicines are available, their prices remain prohibitive for most patients without comprehensive financial protection.

Marked price differences were also observed among hormonal therapies and several cytotoxic medicines. Medicines such as anastrozole, letrozole, docetaxel and oxaliplatin demonstrated several-fold differences between facilities despite being widely used and available from multiple manufacturers. Such variation is unlikely to reflect differences in medicine quality but rather differences in procurement efficiency, purchasing volumes, supplier negotiations and pricing policies. WHO/HAI medicine price surveys consistently demonstrate that decentralized purchasing and weak price regulation contribute to substantial variability in patient prices (19).

Comparison with international reference prices further demonstrated that several generic medicines, including cyclophosphamide, vincristine and filgrastim, were competitively priced, indicating that efficient procurement of selected essential medicines is achievable. However, platinum compounds, including cisplatin and carboplatin, consistently exceeded international reference prices, while prices at the highest-cost facilities frequently surpassed benchmark values by several-fold. These findings suggest that opportunities remain to improve procurement efficiency through pooled purchasing, competitive tendering and stronger price negotiations.

Overall, the observed price variation indicates that patients with the same cancer may face substantially different treatment costs depending solely on the facility where care is obtained. Such variability undermines equity in cancer care and increases the likelihood of catastrophic health expenditure, delayed treatment initiation and treatment interruption. Strengthening national procurement mechanisms, improving price transparency, expanding pooled purchasing and accelerating access to quality-assured biosimilars are likely to reduce price variation and improve affordability of essential cancer medicines in Kenya.

### 4.3 Affordability of Medicines and Chemotherapy Regimens

This study found that the affordability of cancer treatment in Kisumu County remains a major barrier to access. Using the WHO/HAI affordability approach, many anticancer medicines and chemotherapy regimens required several days’ or even months’ wages for the lowest-paid government worker, indicating that without substantive insurance, treatment remains beyond the financial reach of many patients. These findings are consistent with the systematic review by Ocran Mattila et al. (2021), which identified affordability as one of the greatest barriers to accessing cancer medicines across LMICs (22). Similarly, in Ghana and in Rwanda although essential cancer medicines were available, the cost of treatment frequently exceeded patients’ ability to pay, resulting in substantial out-of-pocket expenditure (10, 14).

Affordability varied considerably across cancer types. For breast cancer, conventional chemotherapy regimens were relatively affordable compared with targeted therapies. However, trastuzumab required several months’ wages for a single treatment cycle, making optimal treatment for HER2-positive breast cancer unaffordable for most patients. Cherny et al. (2025) has documented that high out-of-pocket costs remain a major obstacle to accessing innovative cancer therapies worldwide (4). Consistent with our findings, Kizub et al. (2022), reported that trastuzumab required several days’ wages per dose across sub-Saharan African settings (8).

For cervical cancer, weekly cisplatin-based chemotherapy required relatively fewer days’ wages than targeted therapy, suggesting that standard treatment is financially more accessible. However, the affordability of bevacizumab for recurrent or metastatic disease remained extremely poor, limiting access to evidence-based treatment for patients requiring advanced therapy. Likewise, patients with prostate cancer faced a substantial financial burden, particularly for docetaxel-based chemotherapy and newer hormonal agents, which required several weeks’ to months’ wages. These findings support those of Kizub et al. (2022), who reported that access to WHO essential cancer medicines in Kenya, Rwanda and Uganda remains constrained by poor affordability, particularly for high-cost medicines (8).

Although oesophageal and colorectal cancer regimens consisted largely of conventional cytotoxic medicines, the cumulative cost of combination chemotherapy required between one- and four-weeks’ wages for a single treatment cycle. Since treatment typically involves multiple cycles, the overall financial burden becomes considerably greater than that reflected by individual medicine costs. Consistent with our findings, high affordability burdens for colorectal cancer treatment has been documented by Higgins et al. (2026) across sub-Saharan Africa (7). Likewise, the observation is consistent with Lane et al. (2024), who concluded that repeated treatment costs are a major contributor to financial hardship among cancer patients in Africa (9).

Importantly, the affordability estimates presented in this study reflect medicine costs alone and do not include consultation fees, diagnostic investigations, transport, accommodation, medicine administration or income lost during treatment. Consequently, the actual economic burden experienced by patients is likely to be substantially higher. As emphasized by Vogler (2021), improving affordability requires more than reducing medicine prices. It also requires sustainable financing mechanisms that protect patients from catastrophic health expenditure (12). The recent expansion of oncology financing under Kenya’s Social Health Authority provides an important opportunity to improve financial protection. However, continued investment in comprehensive reimbursement of essential cancer medicines and chemotherapy regimens will be necessary to ensure equitable access to cancer treatment.

### 4.4 Implications of All Available Evidence

Taken together, price, procurement governance, and insurance package design determine cancer medicine access. Financial toxicity in cancer care persists even in settings expanding toward universal health coverage, particularly where medicine price regulation and procurement coordination remain weak. The KES 800,000 increase substantially strengthens coverage for standard, fixed-cycle cytotoxic chemotherapy for breast, cervical, oesophageal, and colorectal regimens that already consumed a modest share of the prior cap. However, the revision does not resolve the affordability gap identified for targeted and continuous therapies. More critically, therapies administered continuously rather than over a fixed number of cycles remain poorly served by a single annual cap of any size. Our extrapolation of abiraterone pricing data indicates that this agent alone, taken continuously for metastatic castration-resistant prostate cancer, could consume 75–90% of the enhanced cap within twelve months, independent of any other treatment needs in the same benefit year. These findings indicate that the KES 800,000 revision is a meaningful and welcome policy response to the affordability burden, but not a substitute for the structural redesign of SHA package.

### 4.7 Study Strengths and Limitations

This study is the first systematic, standardized, facility-level oncology medicine survey in Kisumu County and Western Kenya using a standardized WHO/HAI methodology adapted for cancer medicines, allowing comparability with international studies. Critically, the study provides the first evidence-based assessment of the adequacy of Kenya’s SHA KSh 550,000 annual oncology benefit package and the most recently implemented KSH 800,000 benefit package against actual full-course treatment costs. The study demonstrates that access inequity is embedded within the health system itself driven by procurement and regulatory failures, not national economic conditions alone. The study employed UNICEF MPR benchmarking further demonstrating that competitive procurement for core cytotoxics is achievable in this setting, providing a concrete target for pooled procurement reform. However, the small number of surveyed facilities limits statistical generalization. The indirect patient costs such as transportation, diagnostics, lost productivity were not included and would substantially increase the true economic burden. Future research should incorporate longitudinal stock tracking, patient-level cost data, procurement price benchmarking against KEMSA actual costs, and expansion to other Kenyan counties to assess the generalizability of these findings.

## 5. CONCLUSION

Cancer medicines in Kisumu County remain insufficiently available, variably priced, and frequently unaffordable despite insurance expansion. Generic penetration has improved access to conventional cytotoxic medicines but targeted therapies and biologic agents remain largely inaccessible. Inter-facility price variation demonstrates persistent inequities in access driven by fragmented procurement and weak price regulation. Availability alone does not guarantee access. Procurement systems, medicine pricing, and insurance benefit design remain central determinants of whether patients ultimately receive treatment. Strengthening centralized procurement and pooled purchasing could improve medicine availability while reducing inter-facility price variation, at the same time improving access to high-cost oncology medicines. Price markup regulation targeting same-product differentials, KEML expansion for priority targeted therapies, and SHA packages redesigned around full-course regimen costs could minimize out of pocket costs and treatment interruption. Notably, a subsequent SHA oncology cap raised from KES 550,000 to KES 800,000 indicates that this increase improves coverage for standard cytotoxic regimens but does not resolve the affordability gap for targeted therapies and continuous hormonal treatment for disease-specific benefit redesign.

## DATA AVAILABILITY

The data that support this study may be shared upon reasonable request to the corresponding author, if appropriate.

## AUTHOR CONTRIBUTIONS

JOO, SAA, and DOO conceived and designed the study. JOO drafted the manuscript, with revisions and supervisory support from SAA and DOO. All authors read and approved the final manuscript.

## ACKNOWLEDGEMENTS

We thank all study participants for their cooperation. We appreciate the Ministry of Health, County Government of Kisumu for permission to conduct this study. We are grateful to the management and oncology pharmacy staff of health facilities in Kisumu, Kenya, for their support during data collection.

## REFERENCES

1. Sung H, Ferlay J, Siegel RL, Laversanne M, Soerjomataram I, Jemal A, et al. Global Cancer Statistics 2020: GLOBOCAN Estimates of Incidence and Mortality Worldwide for 36 Cancers in 185 Countries. CA Cancer J Clin. 2021;71(3):209–49.

2. GLOBOCAN. Global Cancer Observatory. 2022.

3. Ferlay J, Ervik M, Lam F, Laversanne M, Colombet M, Mery L, et al. Global Cancer Observatory: Cancer Today. Lyon, France: International Agency for Research on Cancer. Available from: https://gco.iarc.who.int/today, accessed [09 09 2024] 2024 [Available from: https://gco.iarc.who.int/today.

4. Cherny NI, Trapani D, Galotti M, Saar M, Bricalli G, Roitberg F, et al. ESMO Global Consortium Study on the availability, out-of-pocket costs, and accessibility of cancer medicines: 2023 update. Ann Oncol. 2025;36(3):247–62.

5. Zhu Y, Wang Y, Sun X, Li X. Availability, Price and Affordability of Anticancer Medicines: Evidence from Two Cross-Sectional Surveys in the Jiangsu Province, China. Int J Environ Res Public Health. 2019;16(19).

6. Wilfinger MJ, Jack Doohan, Ekezie Okorigwe, Ayenew Ashenef, Atalay Mulu Fentie, Ibrahim Chikowe, et al. Substandard anticancer medications in clinical care settings and private pharmacies in sub-Saharan Africa: a systematic pharmaceutical investigation. Lancet Glob Health. 2025;13: e1250–57.

7. Higgins CR, Wilfinger MJ, Doohan J, Okorigwe E, Ashenef A, Fentie AM, et al. Chemotherapy Medications in Sub-Saharan Africa: Availability, Pricing, Affordability, and Predictors of Quality. JCO Glob Oncol. 2026;12:e2500118.

8. Kizub DA, Naik S, Abogan AA, Pain D, Sammut S, Shulman LN, et al. Access to and Affordability of World Health Organization Essential Medicines for Cancer in Sub-Saharan Africa: Examples from Kenya, Rwanda, and Uganda. Oncologist. 2022;27(11):958–70.

9. Lane J, Nakambale H, Kadakia A, Dambisya Y, Stergachis A, Odoch WD. A systematic scoping review of medicine availability and affordability in Africa. BMC Health Serv Res. 2024;24(1):91.

10. Ocran Mattila P, Biritwum RB, Babar ZU. A comprehensive survey of cancer medicines prices, availability and affordability in Ghana. PLoS One. 2023;18(5):e0279817.

11. Petricca K, Carson L, Kambugu J, Denburg A. Strengthening access to cancer medicines for children in East Africa: policy options to enhance medicine procurement, forecasting, and regulations. Glob Health Res Policy. 2024;9(1):24.

12. Vogler S. Can we achieve affordable cancer medicine prices? Developing a pathway for change. Expert Rev Pharmacoecon Outcomes Res. 2021;21(3):321–5.

13. National Cancer Institute Kenya. Significant Boost to Oncology Funding Under SHA: National Cancer Institute Kenya; 2025 [Available from: https://wp.ncikenya.go.ke/2025/04/02/significant-boost-to-oncology-funding-under-sha/.

14. Rurangwa C, Ndayisenga J, Sezirahiga J, Nyirimigabo E. Availability and affordability of anticancer medicines at cancer treating hospitals in Rwanda. BMC Health Serv Res. 2023;23(1):717.

15. Fundytus A, Manju Sengar, Dorothy Lombe, Wilma Hopman, Matthew Jalink, Bishal Gyawali, et al. Access to cancer medicines deemed essential by oncologists in 82 countries: an international, cross-sectional survey. www.thelancetcom/oncology. 2021;22:11.

16. Alemu BA, Hailemariam FH. Price, Availability and Affordability of Anti-Cancer Medicines in Addis Ababa, Ethiopia. Risk Manag Healthc Policy. 2022;15:2421–33.

17. Martei YM, Chiyapo S, Grover S, Ramogola-Masire D, Dryden-Peterson S, Shulman LN, et al. Availability of WHO Essential Medicines for Cancer Treatment in Botswana. J Glob Oncol. 2018;4:1–8.

18. Cherny NI, Sullivan R, Torode J, Saar M, Eniu A. ESMO International Consortium Study on the availability, out-of-pocket costs and accessibility of antineoplastic medicines in countries outside of Europe. Ann Oncol. 2017;28(11):2633–47.

19. WHO. Pricing of cancer medicines and its impacts. Geneva: World Health Organization; 2018. Licence: CC BY-NC-SA 3.0 IGO.; 2018.

20. Joshi D, Khursheed R, Gupta S, Wadhwa D, Singh TG, Sharma S, et al. Biosimilars in Oncology: Latest Trends and Regulatory Status. Pharmaceutics. 2022;14(12).

21. Habashy C, Yemeke TT, Bolous NS, Chen Y, Ozawa S, Bhakta N, et al. Variations in global prices of chemotherapy for childhood cancer: a descriptive analysis. EClinicalMedicine. 2023;60:102005.

22. Ocran Mattila P, Ahmad R, Hasan SS, Babar ZU. Availability, Affordability, Access, and Pricing of Anti-cancer Medicines in Low- and Middle-Income Countries: A Systematic Review of Literature. Front Public Health. 2021;9:628744.

